# From Carb Counting to Diagnosis: Real World Patient Uses and Attitudes Toward Large Language Models in Diabetes Management

**DOI:** 10.64898/2026.03.10.26348079

**Authors:** Raisa Ntemenyi Nkweteyim, Vishesh Girish Shet, Sandra Iregbu, Lu He

## Abstract

Managing diabetes-related conditions is time-intensive and cognitively demanding for patients and caregivers, requiring ongoing glucose monitoring, dietary regulation, physical activity planning, and continuous lifestyle adaptation. With the emergence of large language models (LLMs), patients have increasingly turned to these tools for information, guidance, and support. However, there is limited empirical understanding of which diabetes-related medical tasks patients delegate to LLMs and what their experiences are. To address this gap, we combined qualitative thematic analysis with LLM-assisted analysis to examine patient attitudes and real-world use cases in using LLMs for diabetes-related tasks. Our analysis identified diverse application areas, ranging from clinical interpretation to nutrition and diet support, and disease management amongst others. LLMs functioned not only as information sources, but as interpretive, analytical, decision-support, emotional, and logistical aids supporting patients’ self-management. Last, we discuss implications for integrating LLMs into patients’ self-management support ecosystems and identify areas that require support and safeguards.

## INTRODUCTION

Diabetes and related metabolic conditions represent a substantial and growing public health burden worldwide. More than 40 million Americans have diabetes, accounting for about 1 in 8 adults. Additionally, more than 2 in 5 adults have experienced prediabetes or insulin resistance^1^, conditions that often require ongoing lifestyle modification and medical monitoring to prevent progression and complications. Diabetes is also associated with elevated cardiovascular risk and increased mortality. Adults with diabetes experience a 75% higher risk of all-cause mortality compared with those without diabetes, with cardiovascular disease (CVD) responsible for a large share of this excess risk^2^. Effective diabetes management depends heavily on sustained self-care, including frequent glucose monitoring, dietary planning, physical activity regulation, medication adherence, and interpretation of evolving health data^3^. However, for patients and caregivers, this work is time-intensive, cognitively demanding, and emotionally taxing, particularly as treatment recommendations evolve, and care is distributed across clinical encounters, digital tools, and informal support networks^4^. These challenges are further compounded by disparities in access to diabetes education, specialist care, and timely clinical guidance^5^, leaving patients and caregivers to navigate complex decisions outside formal healthcare settings.

The landscape of diabetes self-management is rapidly evolving with the proliferation of patient-facing digital health technologies, including mobile health (mHealth) applications, wearable devices, telehealth platforms, and, most recently, AI-driven tools^6^. In the United States alone, more than 6,500 diabetes-related mobile applications were available across major app stores 2025, collectively serving over 38 million users who use these tools for tasks such as glucose monitoring, insulin tracking, dietary planning, and lifestyle support^7^. Among these applications, blood glucose tracking tools account for the largest share of use (41%), while AI-enabled predictive applications represent the fastest-growing segment, expanding at an annual rate of 10.4%^7^.

AI-enabled, conversational coaching features are the newest addition to the diabetes technology ecosystem. A systematic review and meta-analysis of chatbots for diabetes self-management found that most chatbots (68%) provide education and management focused on diet, exercise, glucose monitoring, medications, and complications, and that chatbot interventions were effective in lowering HbA1c^8^. Qualitative evaluations show that both patients and specialists value the on-demand availability, personalized advice, and efficiency of generative AI chatbots for daily diabetes management between clinical visits^9^.

Since the release of ChatGPT in late 2022, large language model (LLM) chatbots have increasingly been used for a broader range of health-related tasks, including health information seeking, mental health support, and health self-management^10^. Although traditional sources such as search engines and health websites remain dominant, a web-based survey found that approximately 21% of respondents reported using LLM chatbots such as ChatGPT for health-related information^11^. One emerging application of LLMs is in patient education and engagement. For instance, ChatGPT has been used to answer common questions about diabetes self-management such as diet, exercise, and insulin use, with generally clear and understandable responses^12^. However, LLM recommendations may vary depending on prompting strategies and model versions and may favor newer, more expensive medications without adequately considering cost or patient access^13^. Commentary in *Diabetes Care* highlighted that while models such as GPT-4 show promise in supporting complex diabetes treatment decisions, careful validation and clinician oversight remain essential^14^.

Despite growing interest in the potential of LLMs to support self-management of diabetes, empirical evidence describing how patients use these tools in real-world settings for diabetes management as well as their experiences remains limited^15^. Existing studies of LLM-based diabetes chatbots and clinical decision support systems primarily evaluate system performance metrics such as accuracy or readability, rather than organic, patient-reported use cases and experience^16^. Understanding patients’ real-world use cases and experiences can help identify areas of improvement for general-purpose LLMs for diabetes management and pinpoint potential risks in their current usage. To address this gap, we conducted a mixed-methods analysis of posts discussing LLM use within diabetes-focused online communities. Using a structured qualitative codebook and complementary computational analysis, we characterized user attitudes toward LLM use and common use cases for diabetes-related tasks. Our findings provide early empirical evidence of how patients integrate general-purpose LLMs into diabetes self-management and highlight implications for the safe and effective design of patient-facing AI technologies and integration into broader chronic care ecosystems.

## MATERIAL AND METHODS

### Data Collection

We selected Reddit as our data source due to its sustained popularity among users who post on this platform to exchange information and support across their journeys of managing health conditions^17^. We conducted a pilot search to identify diabetes-related subreddits and included r/diabetes_t2, r/diabetes, r/diabetes_r1, r/Type1Diabetes, r/prediabetes, r/InsulinResistance, r/GestationalDiabetes, r/TandemDiabetes for our study. We decided to include pre-diabetes, insulin resistance, and both Type 1 and Type 2 diabetes due to the similar nature of self-management tasks involved, despite potentially different disease mechanisms. We collected posts that were created between 2023/1/1 to 2026/1/29 using Reddit’s PRAW API and PushShift’s Reddit data archive Dump, since the wide, consumer-facing adoption of LLMs such as ChatGPT started in early 2023. To identify posts that mentioned LLMs, we tested several LLM keywords and after screening, we used the following keywords: LLM, GPT, Gemini, Grok, Copilot, AI, Large Language Model, ChatGPT. We initially used a more comprehensive list of LLM names but during our pilot search, only a few were actually present in the data. We retrieved both original posts and comments that included these keywords. In total, 852 posts and 1,848 comments were retrieved and included for data analysis.

This study analyzed publicly available posts from Reddit discussing the use of LLMs for diabetes-related tasks. The research did not involve interaction with individuals or the collection of identifiable private information or protected health information. All data were obtained from publicly accessible online forums. When presenting illustrative excerpts, quotations were minimally edited where necessary to reduce traceability while preserving their original meaning.

### Data Analysis

#### Qualitative coding

We followed a commonly adopted thematic analysis procedure^18^. During open coding, three coders (RN, LH, VS) independently coded 60 posts to identify emerging themes around use cases and attitudes to construct a codebook, through weekly discussions of codes. The codebook was iteratively refined after coding another 40 posts, with codes merged, renamed, or removed. After the codebook was established, the same three coders independently applied the code to 140 posts (100 posts from codebook construction and 40 new posts) and calculated inter-rater agreement. The agreement ratio for attitude is 0.77 and for use cases is 0.65. Disagreements were resolved through weekly discussions.

#### LLM-assisted Analysis

Due to the number of data (2,700 text entries in total), we could not apply manual coding to all study data. We leveraged advanced LLM models including Gemini (Gemini-2.5-Flash and Gemini-3.1-Pro) and GPT-5 to scale up the codes on the entire study dataset, so that we can examine the prevalence of different use cases and distribution of user attitudes. As the posts were sourced from publicly available social media data and did not include any protected health information, we selected these two models instead of locally deployable models such as llamas, due to computational resource constraints and the consistent evidence of their robust performance across different health-related NLP tasks^19,20^. We iteratively designed and refined prompts to instruct the LLMs to classify each post and comment for three tasks: relevance detection, attitude toward LLM use and use cases. The finalized prompt is provided in our GitHub repository^21^. Model performance was evaluated using established metrics (precision, recall, F1-score) on the 115 manually annotated posts (25 were excluded due to irrelevance after manual review). Due to the small number of posts for the sub-themes in use cases, LLMs showed unsatisfying performance and therefore were not used for this more granular classification task. The model and prompt with the highest performance was subsequently applied to all study data.

## RESULTS

### LLM performance of classifying stances and use cases

The detailed model performance measured by precision, recall, and F1-score is presented in Table 1 below. Overall, Gemini-3.1-Pro outperformed both Gemini-2.5-Flash and GPT-5.3 in most categories. Gemini-3.1-Pro also demonstrated high performance (over 0.75 F1-score), except for Emotional Support, Disease Management, and Medical Device Troubleshooting, and Diabetes Education and Self Learning. All descriptive statistics reported in subsequent sections are based on classification produced by Gemin-3.1-Pro, as it is the highest performing model. In total, the model identified 623 relevant posts and 967 relevant comments, adding up to 1,590 data points.

**Table 1.**
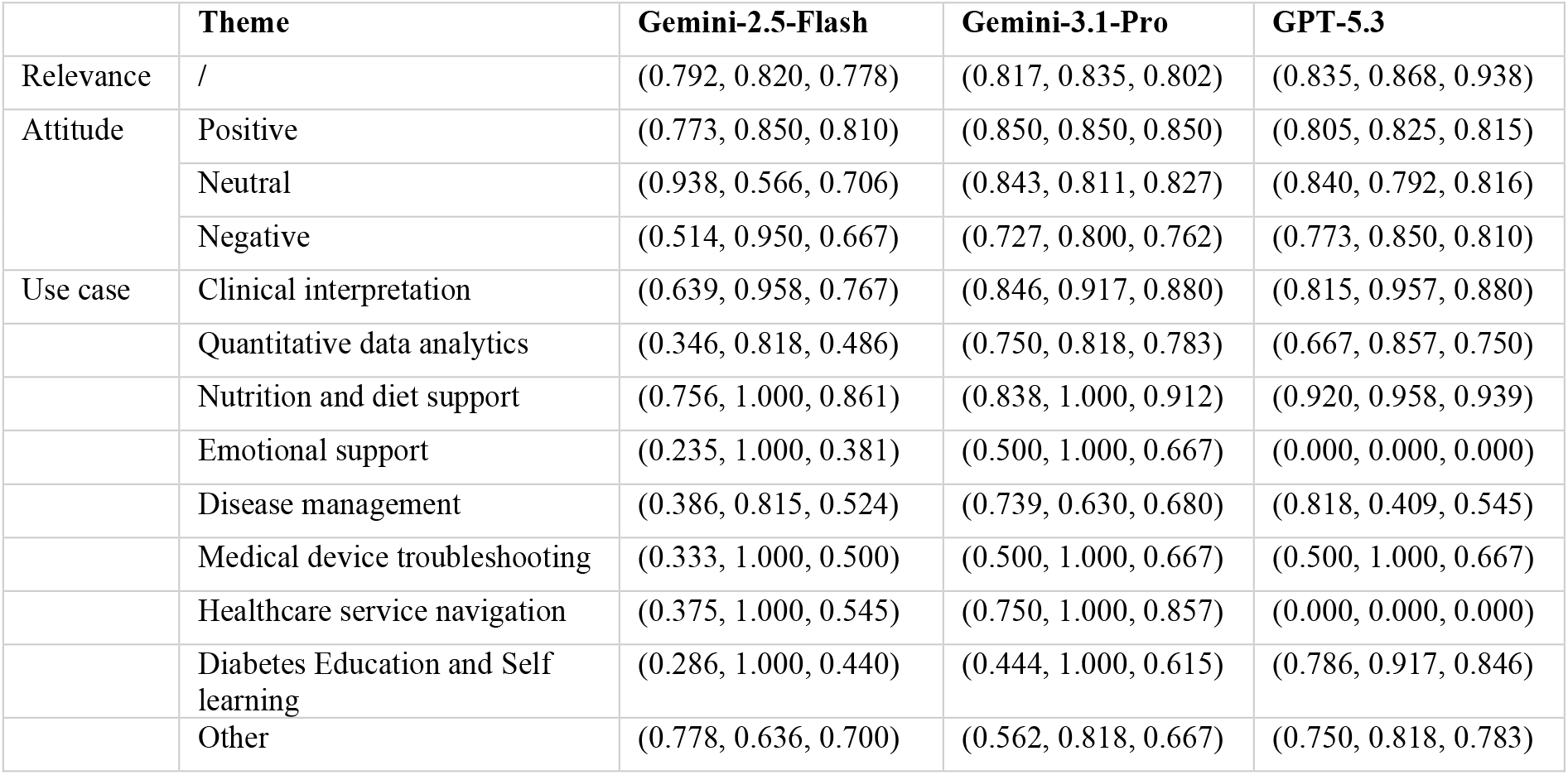
Performance of LLMs for classifying attitude and use cases compared against 115 annotated posts, presented as (Precision, Recall, F1-score)

|  | Theme | Gemini-2.5-Flash | Gemini-3.1-Pro | GPT-5.3 |
| --- | --- | --- | --- | --- |
| Relevance | / | (0.792, 0.820, 0.778) | (0.817, 0.835, 0.802) | (0.835, 0.868, 0.938) |
| Attitude | Positive | (0.773, 0.850, 0.810) | (0.850, 0.850, 0.850) | (0.805, 0.825, 0.815) |
|  | Neutral | (0.938, 0.566, 0.706) | (0.843, 0.811, 0.827) | (0.840, 0.792, 0.816) |
|  | Negative | (0.514, 0.950, 0.667) | (0.727, 0.800, 0.762) | (0.773, 0.850, 0.810) |
| Use case | Clinical interpretation | (0.639, 0.958, 0.767) | (0.846, 0.917, 0.880) | (0.815, 0.957, 0.880) |
|  | Quantitative data analytics | (0.346, 0.818, 0.486) | (0.750, 0.818, 0.783) | (0.667, 0.857, 0.750) |
|  | Nutrition and diet support | (0.756, 1.000, 0.861) | (0.838, 1.000, 0.912) | (0.920, 0.958, 0.939) |
|  | Emotional support | (0.235, 1.000, 0.381) | (0.500, 1.000, 0.667) | (0.000, 0.000, 0.000) |
|  | Disease management | (0.386, 0.815, 0.524) | (0.739, 0.630, 0.680) | (0.818, 0.409, 0.545) |
|  | Medical device troubleshooting | (0.333, 1.000, 0.500) | (0.500, 1.000, 0.667) | (0.500, 1.000, 0.667) |
|  | Healthcare service navigation | (0.375, 1.000, 0.545) | (0.750, 1.000, 0.857) | (0.000, 0.000, 0.000) |
|  | Diabetes Education and Self learning | (0.286, 1.000, 0.440) | (0.444, 1.000, 0.615) | (0.786, 0.917, 0.846) |
|  | Other | (0.778, 0.636, 0.700) | (0.562, 0.818, 0.667) | (0.750, 0.818, 0.783) |

### Attitudes toward using LLMs for diabetes-related tasks

In total, 21% (N = 566) posts and comments expressed positive attitudes describing their experiences using LLMs for diabetes-related, while 19.9% (N = 536) expressed negative attitudes. There are 59.2% (N = 1,598) of the posts that are neutral and did not express any attitudes explicitly. This suggests that, in our study sample, there is no dominant attitude toward LLM use or experiences, with majority being neutral descriptions. When expressing dissatisfaction toward the use of LLMs, users critiqued their lack of ability to provide straightforward answers, or answers that aligned with clinicians’ responses. For example, “*I tried chatGPT for ideas, seemed to want me to eat 6 eggs a day, and suggested foods the nutritionist said to avoid*… *So I think it was good for an idea of what to do, but not reliable at all*.*”* Users also voiced caution toward trusting responses from LLMs, e.g., “*I remember finding discussed in a gastroparesis forum some gel pills that allow oil to coat the undigested food in my stomach, but google is beyond useless for looking up this sort of thing. ChatGPT suggested chlorophyll supplements, but I’m obviously taking that with a salt lick given GPT’s issues*.” Among those that expressed positive attitudes toward LLM use experiences, some commented on their accuracy and the ability to reduce patients’ self-management burden. For example, one user shared *“I am using Chatgpt now for 2 weeks to control my blood sugar, and (in my opinion) it works absolutely fantastic, i fed it with all my information and now it can tell me how many units per carbs, how to prevent hypo and how to get the highs down eaaaasy. Its way less complicated for me now*.*”*

### Use cases

We identified and mapped nine major categories of applications for using LLMs for diabetes-related self-management. These tasks span a wide range across patients’ diabetic journeys, ranging from sensemaking symptoms, interpreting data, diagnosing, managing the disease, to behavioral changes.

#### Nutrition and diet support (N = 713, 25.9%)

Nutrition and diet support were one of the most common and persistent LLM use cases, appearing across diagnosis, treatment initiation, and long-term management stages. Users frequently rely on LLMs to understand dietary restrictions, estimate carbohydrate content, and learn basic nutritional concepts relevant to diabetes control. These interactions typically involve low-risk data inputs such as food descriptions or general dietary questions. As users transitioned into ongoing self-management, nutrition-related LLM use evolved toward optimization and maintenance, including refining meal plans, balancing dietary preferences with glucose control goals, and sustaining weight or glycemic improvements over time.

Several users described using LLMs to generate diabetes-friendly meal ideas. For example, one user noted, *“I’ve been using ChatGPT to suggest better meals (I had been using it before to meal plan and help control food costs… something I know how to do from hospitality coursework), and it knows to suggest diabetic-friendly options*.*”* Other users reported using LLMs to assist with carbohydrate estimation. One individual explained, *“I don’t know if anyone else has checked out the new Bing ai powered search but it seems like it will make carb count lookups much easier. Instead of looking up each item one at a time you can just give it the list of food you have and calculate the measurement in a fraction of the time*.*”*

#### Disease management (N = 621, 22.5%)

Disease management refers to instances in which individuals used LLMs to obtain practical guidance for managing day-to-day aspects of living with diabetes, including behavior changes and medication and insulin adjustments. In these interactions, users described consulting LLMs for strategies related to lifestyle adjustments, symptom management, or general self-management practices that could help maintain stable glucose levels or address diabetes-related complications. These interactions often occurred during ongoing disease management, when individuals were navigating practical challenges associated with daily care. Users reported asking LLMs for advice about routines, diet, exercise, or other behavioral strategies that might support blood glucose control.

For example, one user shared an LLM generated general advice for managing blood glucose levels, including maintaining routines and balancing daily activities: *“Certainly! Here are some real-life tips for balancing blood sugar levels for individuals with type 1 diabetes: Establish a routine: Try to maintain a consistent daily routine for meals, exercise, and insulin administration. This helps to regulate blood sugar levels more effectively and reduces the likelihood of extreme fluctuation”*. One individual also asked whether there were any precautions associated with obtaining a tattoo while living with diabetes.

In some instances, users described providing the models with detailed personal information, including carbohydrate intake, blood glucose values, and time-of-day data, and requesting optimization of insulin management parameters such as carbohydrate ratios, correction factors, or insulin dosage recommendations. One user described providing her daughter’s glucose monitoring graph to GPT for pump setting suggestion: *“*…*I submitted the screen shot and the question if it could suggest mobi pump setting based off the chart. It also suggested time block basal segments along with carb and ISF time block ratios that I did not include in this post I find this interesting and maybe a tool to use for new users to get dialed in*.*”* These examples illustrate how individuals used LLMs as accessible sources of practical guidance for managing everyday aspects of diabetes care.

#### Diabetes education and self-learning (N = 503, 18.3%)

Self-learning refers to the use of LLMs to acquire general knowledge about diabetes, its mechanisms, treatments, or related health concepts. This included explaining diabetes-related research and recent progresses, physiological mechanisms, and emerging treatment options. Such use was especially prominent among users seeking deeper understanding beyond what they reported receiving in clinical encounters. These exchanges often occurred when users were unable to find clear or relevant information through traditional search engines or when they sought simplified explanations of complex medical issues. For example, one user described learning about the disease itself through chatting with LLMs: *“Ever since I was diagnosed, I’ve started taking an interest in this disease. At first, I learned the basic concepts about diet and metabolism — glycemic index and load, the role of insulin, liver glycogen, the pancreas and beta cells, and so on. But the more I dig in, mostly by talking with Copilot and ChatGPT the more I realize how shallow my knowledge really is*.*”*

#### Quantitative and longitudinal Patient Generated Health Data (PGHD) Analysis (N = 358, 13%)

Quantitative data analysis refers to the use of LLMs to analyze, explain, or derive insights from PGHD, such as glucose monitoring data, diet logs, or physical activity records that are often longitudinal and continuously collected from everyday settings. Users described or directly provided raw continuous glucose monitoring (CGM) data, insulin delivery information, sleep data, or longitudinal glucose summaries to LLMs to identify patterns, create visualization or charts, understand variability, or contextualize lifestyle effects on glucose control. Quantitative analysis of PGHD involved more sensitive, personal health data and greater delegation of analytical reasoning to LLMs. Users often positioned LLMs as tools for summarizing and analyzing trends across multiple data streams, enabling faster interpretation of complex health information that is difficult for manual review and sensemaking. For example, one user described using an LLM to track and analyze glucose and sleep data: *“I’ve started using ChatGPT to log the BG reading and my sleep pattern (from my Apple Watch). I’ve found chatGPT is extremely helpful. It creates charts and tables and helps analyzing data so quickly and accurately*.*”*

#### Clinical Interpretation (N = 314, 11.4%)

Clinical interpretation involves asking an LLM to explain, contextualize, or evaluate the meaning and significance of clinical measurements, diagnostic results, or self-reported symptoms. Prior to formal diagnosis, users primarily engage in symptom checking, using LLMs to contextualize bodily experiences (e.g., sweating, excessive urination, shortened sleep), explore possible explanations, and reduce uncertainty about whether symptoms might be diabetes related. These interactions were typically exploratory and framed in tentative language, reflecting interpretive rather than diagnostic intent. For example, one user described consulting an LLM after experiencing several symptoms and reported that the model suggested diabetes was unlikely: *“I had a really bad headache one day, and late at night with this headache I had blurry vision. The blurry vision was only for a few seconds. A day later I have the sudden urge to always urinate but that seems to be gone after one day*… *I told ChatGPT my symptoms and it said that my chances are pretty low since the symptoms kinda just disappeared and I don’t have any other symptoms*.*”*

Around the point of diagnosis, clinical interpretation shifted toward sensemaking and confirmation, including interpreting laboratory results (e.g., HbA1c values) or to evaluate whether symptoms aligned with a potential diabetes diagnosis. One user described consulting an LLM when evaluating symptoms in a young child: *“We took my 2*.*5 year old to see his pediatrician Friday morning with concerns of him requesting an abnormal amount of fluid and wetting through his overnight diapers even though he’s potty trained. Of course, we consulted ChatGPT and were alarmed that it was so definitively pointing to type 1 diabetes*.*”* In some cases, LLMs produced different judgements from clinicians when interpreting lab results, and users sought peers’ opinions: *“i got my blood sugar and Cholestrol tested, I know the cholesterol is bad but the sugar is the one Iâ€™m confused about. The pharmacist said my sugar is at an okay level but Google and ChatGPT say otherwise*..*can someone explain please?”*

In some cases, users reported using LLMs to seek a second opinion on diagnosis. In these instances, users provided more structured clinical data generated from lab and blood work tests and increasingly positioned LLMs as interpretive support, particularly when clinical explanations from healthcare professionals were perceived as insufficient. For example, one individual reported asking an LLM whether their clinical profile might indicate a different diabetes subtype: *“Around June of 2023 it was early in the morning, and I decided to express my questions regarding my diagnosis to ChatGPT. Based on my family history and lack of antibodies is there anything else that I could have besides type 1? The first thing that GPT mentioned was MODY*.*”*

#### Emotional support (N = 63, 2.3%)

Emotional support refers to instances in which individuals interacted with LLMs to express feelings about living with diabetes, seeking encouragement, or engaging in reflective conversations about their conditions. In several cases, users interacted with LLMs to improve their mood by requesting humor in conversations, while others used these interactions to share personal experiences and reflect on their progress in managing their condition. For some individuals, these conversations appeared to become habitual, suggesting that LLMs may function as a recurring outlet for emotional expression and reflection.

In one instance, a user described discussing treatment options with an LLM and reported becoming emotionally overwhelmed when the model asked: *“What would be the first thing you would do if you were cured?”* The individual responded that they would eat an entire pizza, noting that the exchange prompted a moment of introspection and renewed hope regarding the possibility of improved diabetes condition.

#### Healthcare service navigation (N = 38, 1.4%)

Healthcare service navigation refers to instances in which individuals used LLMs to obtain information about healthcare systems, insurance coverage, medical services, or access to diabetes-related care. This use case represented a lower intensity use case where users reported using LLMs to inquire about insurance coverage for diabetes-related medical devices within the healthcare systems of the countries in which they resided or intended to relocate. For example, one user wrote: *“I’m an EU citizen, and I’m planning to move to Spain soon. I can’t find anywhere (and I tried chat GPT) if Dexcom G6 and t;slim X2 are covered by government health insurance. Is there someone here who knows?”*

#### Medical device troubleshooting (N = 35, 1.3%)

Medical device troubleshooting refers to instances in which individuals consulted LLMs to diagnose or resolve problems related to diabetes management technologies or devices. Some users expressed frustration with diabetes management technologies, particularly continuous glucose monitoring (CGM) devices, and sought assistance from LLMs in troubleshooting potential device-related issues. Reported concerns included frequent or persistent alarms, faulty sensors, and perceived inaccuracies in glucose readings when compared with finger-prick blood glucose tests. For example, one user described attempting to resolve persistent CGM alerts by consulting both online searches and an LLM: *“Yes I’ve googled and asked ChatGPT but nothing is working. My Dexcom has decided to beep every 15 minutes, no highs, no lows, just alarming. Does anyone know how to fix this? I did a silence all for six hours and still the alert every 15 or so minutes!”*.

## DISCUSSION

In this mixed-methods study investigating how patients and caregivers use LLMs for diabetes management in real-world settings, we identified a broad spectrum of use cases ranging from low-intensity informational support to high-intensity therapeutic delegation. Our study serves as one of the first studies to provide rich empirical insights into real-world uses of LLMs in patients’ diabetes-related management. We discuss the implications of our study findings for safe and responsible integration of LLMs into real-world patients’ self-management ecosystems.

### The role of LLMs in patients’ diabetes self-management across disease trajectory and meta-tasks

We conceptualized how LLM use for diabetes management is distributed across different meta-tasks across patients’ disease trajectories, shown in Figure 1. Importantly, use cases varied along a continuum of support and risk intensity at different stages. At the lower end were logistical, informational and educational uses (e.g., troubleshooting medical devices, learning about insurance coverage). At the higher end were cases involving behavioral suggestions including insulin dosing decisions and guidance during acute glycemic events. This gradient highlight that LLM engagement in diabetes management is not monolithic but spans multiple levels of autonomy and delegation, aligning with recent studies on LLM use for health^22^.

**Figure 1.**
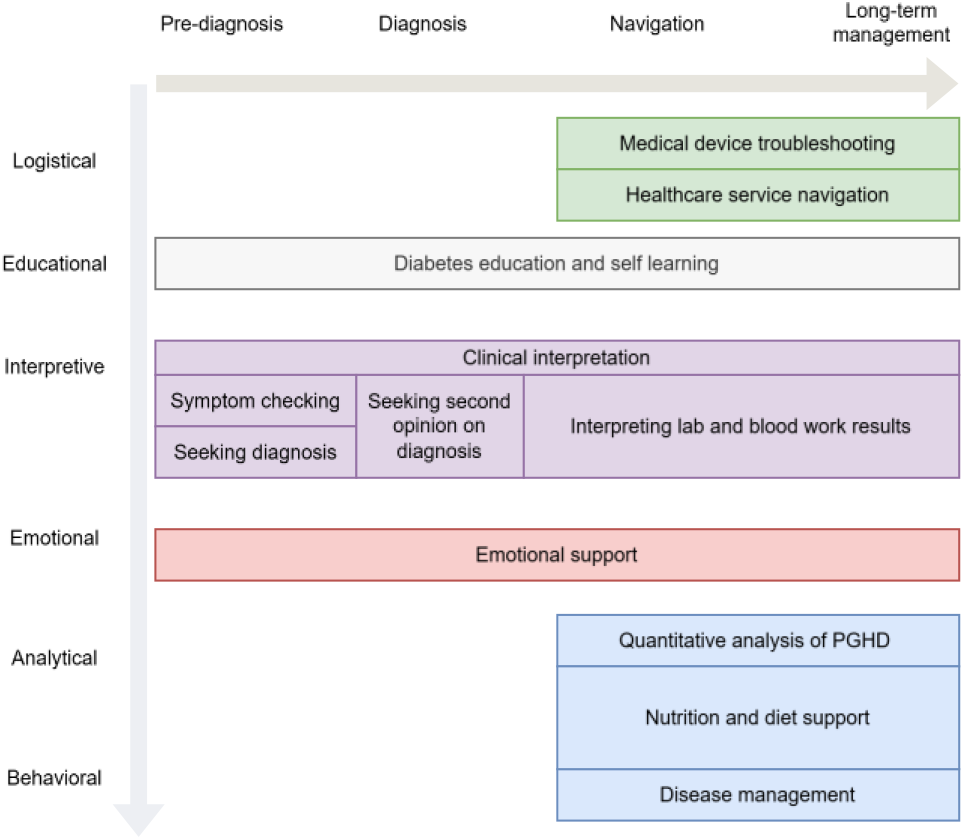
Conceptual mapping of LLM use cases across disease trajectories and for different meta-tasks

We note that this conceptualization was developed based on our analysis and abstraction of the data and is not an absolute conceptual mapping. For example, while patients commonly start tracking PGHD and using them to seek analytical insights from LLMs after diagnosis and during long-term management, some patients may already start doing this before diagnosis. The goal of this mapping is to provide conceptual clarity of the common use case distribution, to inform future research, patient education, and LLM improvement for different disease stages and meta-tasks. We encourage future research to continuously generate empirical evidence to refine and extend our conceptual mapping.

### Potential risks of patients’ use of LLMs for diabetes management

While many reported uses in this study were low risk (e.g., meal brainstorming, macros education), our analysis also identified higher-risk scenarios involving therapeutic delegation. Several risk domains emerged: (1) Therapeutic delegation: users described adjusting insulin regimens or basal profiles based on LLM recommendations. Even when framed cautiously, this represents a transfer of clinical reasoning to a non-regulated system, especially without oversight and confirmation from clinicians. (2) Diagnostic framing: In pre-diagnostic contexts, LLM outputs shaped initial interpretations of symptoms, potentially influencing emotional responses and urgency perceptions. (3) Sensitive data disclosure: Users reported uploading detailed laboratory panels and PGHD into LLM interfaces, raising privacy and data governance concerns. Generic LLM systems lack sufficient safeguards or regulatory compliance^23^ (e.g., Health Insurance Portability and Accountability Act - HIPAA^24^), increasing the risk of confidentiality breaches and misinterpretation due to missing clinical context.

In addition, prior evaluations of LLMs in diabetes care have documented errors such as incorrect information about insulin storage, confusion between insulin regimens, improper guidance on insulin mixing, and misinterpretation of blood glucose units^25^. In high-stakes contexts such as insulin dosing or symptom triage, these inaccuracies could lead to harmful outcomes, including inappropriate dosing or delays in seeking clinical care.

### Responsible integration and regulation

Given the expanding role of LLMs in diabetes self-management, we propose that responsible integration requires a multi-level approach. First, at the technology level, LLMs intended for health contexts should incorporate explicit uncertainty communication and clearly delineate the boundaries between educational information and medical advice. In addition, context-aware guardrails could be particularly important for insulin and medication-related queries. Second, at the clinical practice level, as LLM use and exploration is increasing among patients, rather than prohibiting LLM use, clinicians may need to anticipate and discuss it through open dialogue about how patients are using AI tools. Integration strategies might include co-reviewing AI-generated plans or clarifying safe parameters for self-adjustment. Third, at the policy level, when LLM outputs directly influence insulin dosing or CGM data interpretation, they begin to approach the functional territory of regulated medical devices. Policymakers may therefore need to clarify when AI systems cross into clinical decision support, what validation standards should apply, and how liability should be distributed. Differentiating low-risk informational uses from high-risk therapeutic delegation will be essential to avoid overregulating benign educational applications while still addressing clinically consequential ones. The American Diabetes Association recommends that patients use digital health tools that have undergone FDA review and comply with HIPAA security standards^26^. However, evidence supporting the safety and effectiveness of many AI-driven health applications remains limited^27^. Additional research, particularly well-designed randomized controlled trials, is needed to evaluate their clinical benefits and ensure adequate security protections.

### Implications for patient education

Our findings highlight the importance of strengthening digital health literacy within diabetes education programs. Many users demonstrated awareness that LLM outputs may be incomplete or unreliable, often framing their use cautiously (e.g., “*if ChatGPT is to be believed*”). However, other posts described situations in which inaccurate or oversimplified information could potentially influence health decisions, such as carbohydrate estimation or insulin-related calculations. These observations suggest that patient education should incorporate guidance on appropriate uses of generative AI tools, including how to verify information, recognize limitations, and avoid relying on LLMs for high-risk clinical decisions. Diabetes education should equip patients and caregivers to understand the probabilistic nature of LLM outputs, the distinction between pattern explanation and medical authorization, and the importance of clinician confirmation before making medication changes^28^. Healthcare providers and diabetes educators may consider addressing generative AI explicitly during patient education. Rather than discouraging LLM use entirely which may be impractical given their widespread accessibility, clinicians and educators can help patients understand how to use these tools safely. Providing clear boundaries for safe use could mitigate potential harms while preserving the educational benefits these tools seem to offer.

Finally, the results indicate an opportunity for future digital health interventions to integrate LLM-based educational support into structured diabetes self-management programs. If appropriately designed and clinically validated, conversational AI systems could potentially augment traditional education by offering on-demand explanations of diabetes concepts, lifestyle guidance, and personalized educational reinforcement. However, such integration will require careful attention to safety, transparency, and clinical oversight to ensure that patient-facing AI tools support, not replace, evidence-based diabetes care.

### Limitations and future work

This study has several limitations. First, the analysis relied on publicly available posts from Reddit, which may not represent the broader population of individuals living with diabetes. Users of online forums are often more technologically engaged and may differ from patients who do not participate in digital communities^29^. Second, the findings are based on self-reported posts that could not be independently verified, and many posts lacked contextual information about users’ clinical histories, treatment regimens, or interactions with healthcare providers. Third, the analysis reflects behaviors observed within a specific online community and time period during which LLM technologies were rapidly evolving. As such, the patterns observed in this study may change as models, interfaces, and user familiarity continue to develop. Despite these limitations, the study provides early insight into how individuals are incorporating LLMs into everyday diabetes self-management practices in real-world online environments.

Future work should extend to more in-depth investigation of patients’ and caregivers’ routine use of LLMs for diabetes-related tasks, through interviews, focus groups, and longitudinal diary studies. Studies should probe further to understand how the use LLMs for diabetes management affect patient-provider communication, patients’ skills and burden of self-management, and test various design prototypes to integrate LLMs more organically into their existing management workflows.

## CONCLUSION

Our study provides one of the first empirical evidence of how patients and caregivers use LLMs for diabetes-related tasks in everyday settings. The use cases span a wide spectrum ranging from low-intensity support such as diabetes education to high-intensity tasks including clinical interpretation. Our findings highlight both the promise and the complexity of this emerging support technology: LLMs may enhance self-efficacy and reduce patients’ burden but also introduce new forms of therapeutic delegation and privacy exposure. The central challenge moving forward is not whether patients will use LLMs for diabetes management, but how healthcare systems, designers, and regulators can ensure that such use enhances safety, equity, and clinical integrity rather than undermining them.

## Data Availability

Data will be provided at reasonable request to the author.

